# Pneumonia in Sudan; Systematic Scoping Review of Literature and Meta Analysis

**DOI:** 10.1101/2022.09.23.22280257

**Authors:** D M Mohamed, M A SalahEldin, A B Idris, E B Idris, S G Mohamed, M M Badawi

## Abstract

In addition to excessive burden of non-communicable diseases, natural and manmade disasters and internal conflicts, Sudan is predominantly susceptible to communicable diseases such as Malaria, Tuberculosis, and Pneumonia, which bring about a burden of infectious diseases and demand for high quality health care. According to the WHO as well as the Sudan Health Observatory, Pneumonia is one of the leading causes of death in Sudan. This study therefore aimed to determine pneumonia infection prevalence among Sudanese as well as its related risk factors. A systematic review of the literature was conducted in the 1^st^ of December 2020. The review was regulated in accordance with PRISMA. After abstract and full text screening only seventeen articles met our inclusion criteria and passed the quality assessment procedure. Seven included studies determined prevalence of pneumonia; the overall pooled prevalence was around 30%. Furthermore, twelve research articles investigated risk factors related to pneumonia among Sudanese population. Further research with larger sample sizes targeting risk factors of pneumonia among Sudanese population is needed to be conducted.

## 1 Introduction

In addition to excessive burden of non-communicable diseases, natural and manmade disasters and internal conflicts, Sudan is predominantly susceptible to communicable diseases such as Malaria, Tuberculosis, and Pneumonia, which bring about a burden of infectious diseases and demand for high quality health care^[1]^.

According to the *WHO* as well as the Sudan Health Observatory in the federal ministry of health, the major communicable diseases contributing to morbidity are Malaria, Schistosomiasis, Pneumonia, Tubercelosis and Diarrheal diseases ^[2–4]^.

Pneumonia is the single largest infectious cause of death in children worldwide. It killed 808,694 children under the age of five in 2017, accounting for 15% of all deaths of children under five years old. Pneumonia affects children and families everywhere, but is most prevalent in South Asia and sub-Saharan Africa. According to Sudan Health Observatory; it was the third cause of death in 2017 among inpatients in Sudan as 6% of the total deaths were due to pneumonia ^[5–7]^.

This study therefore aimed to determine what have been done regarding pneumonia infection prevalence among Sudanese as well as its related risk factors, identification of knowledge gaps will be a giant leap and of great value to the health security of human of Sudan.

## 2 Materials and Methods

### 2.1 Search strategy

To identify relevant studies; a systematic review of the literature was conducted in the 1^st^ of June 2022. The review was regulated in accordance with PRISMA-ScR (Preferred Reporting Items for Systematic Reviews and Meta-Analyses extension for Scoping Reviews) ^[8]^ (S1 Table). A comprehensive search was operated in PubMed, Embase, Google scholar, Scopus, Index Copernicus, DOAJ, EBSCO-CINAHL, Cochrane databases without language limits (studies written in languages other than English were later excluded). To obtain a current situation evidence; only studies published in or after 2010 were included. Furthermore, all studies where the data collection process took place before 2010 were also excluded, the only exception was if the collection process started in or before 2010 and ended in 2010 or afterwards.

As medical literature in Sudan is generally scarce in international databases as well as risk factors may be differently reported, risk factors were not used in keyword formulation and their related results were later extracted from included studies. Moreover, studies toward *COVID-19* infection were not included as the current situation is intended to be evaluated despite of the *COVID-19* pandemic; however, studies concerned with *COVID-19* prevalence not risk factors were included. Nevertheless, studies investigating *COVID-19* associated risk factors were cited without their results being scoped to guide readers.

The keywords used in PubMed were as follow:

(Lower respiratory tract infection) OR Pneumonia OR Pneumonias OR (Lung inflammation) OR Lobitis OR (non specific inflammatory lung diseases) OR Prepneumonia OR pleuropneumonia OR Pleuropneumonitis OR (Pneumonic lung) OR (Pneumonic pleurisy) OR (Pneumonic pleuritis) OR Pnemonitides OR Pneumonitis OR (Pulmonal inflammation) OR (Pulmonary inflammation) OR (Pulmonic inflammation) AND Sudan*[tiab]. As previously described ^[9]^

Moreover, to optimize our search, hand searches of reference lists of included articles were also performed.

### 2.2 Study selection and data extraction

Titles and abstracts were assessed for preliminary eligibility. A copy of the full text was obtained for all research articles that were available and approved in principle to be included. Abstraction of data was in accordance with the task separation method; method and result sections in each study were separately abstracted in different occasions to reduce bias. Moreover, data abstraction was conducted with no consideration of author’s qualifications or expertise as described in details previously^[10]^. Each research article was screened for all relevant information and recorded in the data extraction file (Microsoft Excel), data from each method section was extracted using a predefined set of variables; study characteristics, type of the communicable disease, type of participants, study population size, geographical region, methodology used in prevalence or risk assessment and the period of the study conduction.

After inclusion, studies were further classified in each communicable disease into studies determining prevalence, studies determining risk factors and studies determining both prevalence and risk factors. Furthermore, as risk factors-related keywords were not formulated in the search strategy, each study was fully screened to check the nature of the risk investigated by authors, studies in which socio-cultural risks have not been assessed, were later excluded.

When studies included regarding socio-cultural risks of a given communicable disease is scarce where Meta analysis cannot be synthesized, scoping of the literature was conducted to highlight methodologies and results conducted as well as to illustrate gaps of research among Sudanese population.

### 2.3 Assessment of quality

Each included article was evaluated based on a framework for making a summary assessment of the quality. The related published literature was crossed, then a framework was structured specifically to determine the level of representativeness of the studied population and to judge the strength of the estimates provided. Five questions were to be answered in each article, each answer represent either 1 score for yes, 0 score for No or 0 score for not available; a total score for risk of bias and quality was calculated by adding up the scores in all five domains, resulting in a score of between 0 and 5. The highest score indicates the highest quality, only studies with a score for quality greater or equal to 3 (higher quality) were included. The five domains were: is the study objective clearly defined?, is the study sample completely determined?, is the study population clearly defined and specified?, is the methodology rigorous? And is the data analysis rigorous?

### 2.4 Quantitative analysis

Meta-analysis was performed—whenever possible using Review Manager Software (Version 5.3). The software automatically provided the Confidence Interval (CI) according to the calculated *SE*, if the *CI* is provided in a study; it was introduced accordingly. The heterogeneity of each meta-analysis was assessed as well, the random effect was favored over the fixed effect model in all meta-analysis established as variations between studies is predicted to be probable due to the diversity of the study populations. Sensitivity analysis was also approached to determine the effect of studies conducted in populations proposed to behave in indifference manners or proposed to be more aware on the overall pooled prevalence. Moreover, subgroup analysis was also conducted -whenever suitable to determine prevalence or risk level in specific State or population. An outcome to take part in the meta-analysis has to be included in at least two studies.

Trim and Fill method was used to assess the risk of publication bias in each Meta analysis conducted ^[11]^.

## 3 Results

### 3.1 Studies included

A total of 1,420 articles were identified from the search strategy including hand searches of reference lists of included original research articles and reviews. From these, 1,348 articles were excluded. After abstract and full text screening only fifteen articles met our inclusion criteria and passed the quality assessment procedure. The articles reported prevalence among specific population and/or risk factors. (Fig. 1) illustrates the PRISMA flow diagram. The included articles are depicted in (Table 1). Assessment of the quality of included studies is depicted in (Table S2).

**(Table 1):** Characteristics of included studies

| **Study ID** | **Year of publication** | **Study design** | **City/State** | **study population/s** | **Assessment of** | **sample size** | **Gender** | **Participants' Age** |
| --- | --- | --- | --- | --- | --- | --- | --- | --- |
| Abd Elrhman E.,2015 [<sup>12</sup>] | 2015 | Cross sectional | Khartoum State | Suspected Patients | Prevalence of M. Catarrhalis (Bacteriological methods) | 200 | Both | 19-86 years |
| Essa M.,2017 [<sup>13</sup>] | 2017 | Retrospective | South Darfur State | Suspected Patients | Prevalence | 9000 | Both | 1-44 years |
| Gabbad A.,2014 <sup>14</sup> | 2014 | Cross Sectional | Omdurman City | Children | Prevalence of S. pneumoniae (Bacteriological methods) & Risk factor | 282 | Both | < 5 years |
| Gritly S.,2018 [<sup>15</sup>] | 2018 | Cross Sectional | Omdurman City | Children | Risk Factors | 40 | Both | < 5 years |
| Idress W.,2017 [<sup>16</sup>] | 2017 | Cross Sectional | Khartoum State | Nurses | Risk Factors | 60 | Both | 20 - > 40 years |
| Mahgoub A.,2013 [<sup>17</sup>] | 2013 | Cross Sectional | Khartoum State | Children | Prevalence & Risk Factors | 260 | Not determined | Not determined |
| Mohammed S.,2013 [<sup>18</sup>] | 2013 | Cross Sectional | Gezira State | Mothers of children less than 5 years | Risk Factors | 250 | Female | 18-63 years |
| Mohamoud A.,2014 [<sup>19</sup>] | 2014 | Cross Sectional | Gezira State | Parents of children less than 5 years | Risk Factors | 182 | Both | 20 - > 50 years |
| Moser W.,2022 [<sup>20</sup>] | 2022 | Cross Sectional | Omdurman City | General population | Prevalence of SARS-Cov-2 (ELISA) & Risk factor | 2374 | Not determined | < 5 - ≥ 50 years |
| Omar M.,2016 [<sup>21</sup>] | 2016 | Cross sectional | Kassala City | Pulmonary TB Patients | Prevalence of S. pneumoniae (Bacteriological methods) | 150 | Both | 10-50 years |
| Salah E.,2015 [<sup>22</sup>] | 2015 | Cross Sectional | Omdurman City | Children with pneumonia | Risk Factors | 150 | Both | 2-60 months |
| Salih K.,2011 [<sup>23</sup>] | 2011 | Cross Sectional | Khartoum State | Children with severe pneumonia | Risk Factors | 224 | Both | 0-59 months |
| Salih K.,2014 [<sup>24</sup>] | 2014 | Cross Sectional | Khartoum State | Children with severe pneumonia | Risk Factors | 208 | Both | mean 28 months |
| Salih K.,2015 [<sup>25</sup>] | 2015 | Cross Sectional | Khartoum State | Children with severe pneumonia | Risk Factors | 195 | Both | 2-59 months |
| Salih K.,2015 [<sup>26</sup>] | 2015 | Cross Sectional | Khartoum State | Children with severe pneumonia symptoms | Prevalence ( Bacteriological Methods) | 189 | Both | < 5 years |

**Fig (1):**
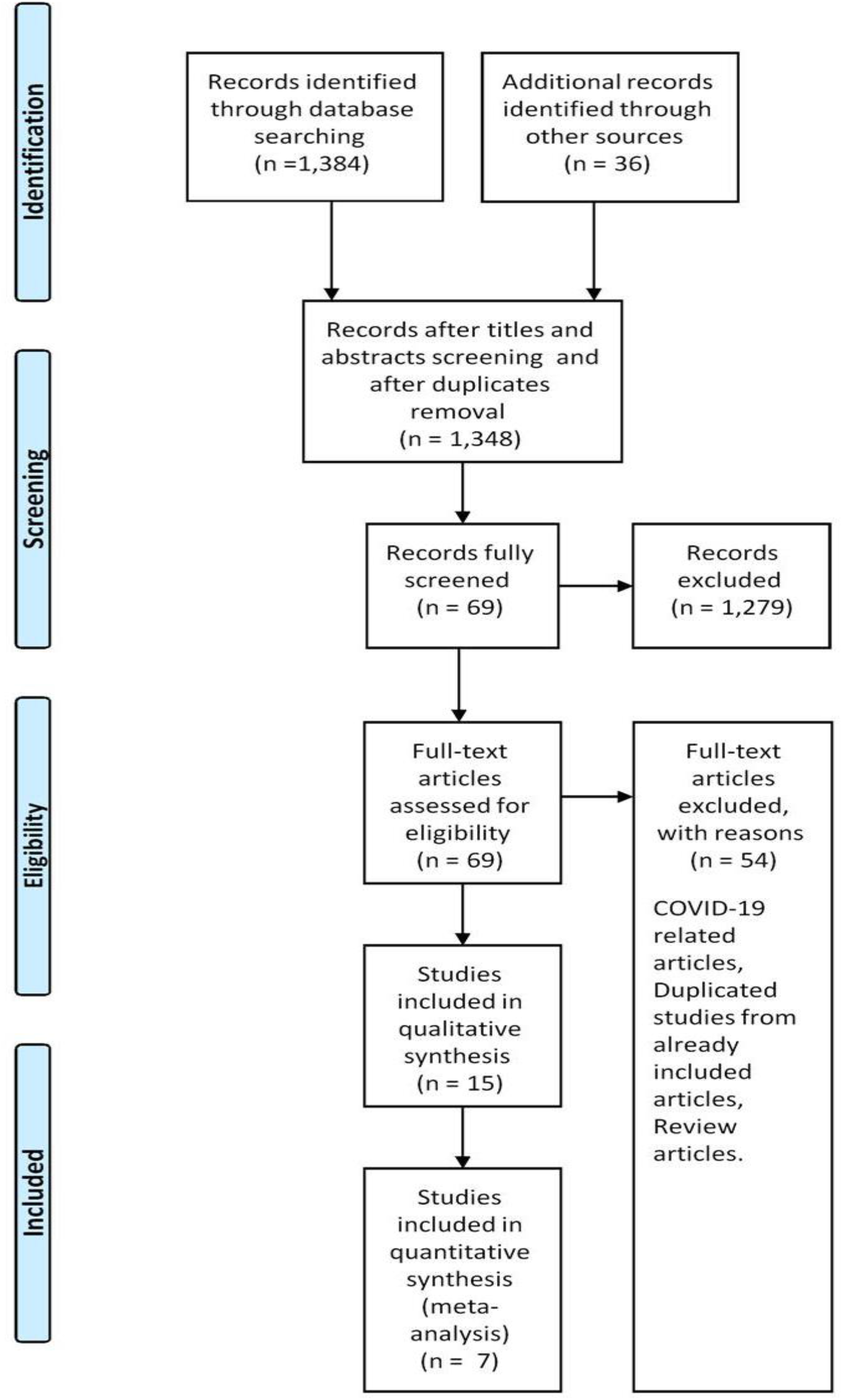
PRISMA flow diagram

### 3.2 Study characteristics

The characteristics of the included studies are depicted in (Table 1), among which the oldest were published in 2011 ^[23]^ while the most recent one was published in 2022 ^[20]^. Eleven research articles determining pneumonia prevalence or risk factors conducted in Khartoum State ^[12,14–17,20–26]^, two in Gezira State ^[18,19]^ and one in South Darfur State ^[13]^. Moreover, almost all articles were conducted among both gender, nevertheless, one study recruited females only (mothers of infected children) ^[18]^ while another study did not determine the gender of their participants ^]17]^.

Furthermore, 10 articles were concerned of pneumonia prevalence/risk factors among children ^[14,15,17–19,22–26]^. All characteristics of included studies are depicted in (Table 1). Publication bias assessment indicated no major asymmetry.

### 3.3 COVID 19 Studies

Studies concerned with pathogenesis, health services, knowledge, behavior or risks related to *COVID-19* were beyond the scope of this review. Nevertheless, to guide interested readers as well as to pool literature related to pneumonia among Sudanese population and for further research prospect, all related articles according to the current search strategy are depicted in (Table 2).

**Table (2).** Studies toward *COVID-19* among Sudanese population according to search strategy

| Author | Year of publication | Title |
| --- | --- | --- |
| Abd El-Raheem et al [27] | 2022 | Extent of COVID-19 Healthcare Services of Isolation Center of Private Hospital across Khartoum State, Sudan |
| Abd El-Raheem et al [28] | 2021 | Characteristics and severity of COVID-19 among Sudanese patients during the waves of the pandemic |
| Abdelmagid et al [29] | 2021 | Acceptability and feasibility of strategies to shield the vulnerable during the COVID-19 outbreak: a qualitative study in six Sudanese communities |
| Abdou et al [30] | 2021 | The coronavirus disease 2019 (COVID-19) vaccination psychological antecedent assessment using the Arabic 5c validated tool: An online survey in 13 Arab countries |
| Abouzid et al [31] | 2021 | Influence of COVID-19 on lifestyle behaviors in the Middle East and North Africa Region: a survey of 5896 individuals |
| Ahmed et al [32] | 2021 | Impact of the COVID-19 pandemic on intimate partner violence in Sudan, Malawi and Kenya |
| Ahmed et al [33] | 2021 | COVID-19 in Sudan |
| Ahmed et al [34] | 2022 | Clinical characteristics, complications, and predictors of outcome of hospitalized adult Sudanese patients with COVID-19 and malaria coinfection in Sudan: A multicenter retrospective cross-sectional study |
| Ahmed et al [35] | 2020 | Controlling the spread of COVID-19 in Sudan with limited resources: a unique community-engaged approach |
| Ali et al [36] | 2022 | The role of integrated programs in the prevention of COVID-19 in a humanitarian setting |
| Aljak et al [37] | 2021 | The first laboratory-confirmed imported infections of SARS-CoV-2 in Sudan |
| Attaallah et al [38] | 2021 | The Prognostic Value of Thrombocytopenia in COVID-19 Pneumonia, Gezira Isolation Centers, Sudan |
| Aung et al [39] | 2021 | Community responses to COVID-19 pandemic first wave containment measures: a multinational study |
| Bara'a et al [40] | 2021 | Coronavirus 2019-like illness and public adherence to preventive measures, Sudan 2020 |
| Deek et al [41] | 2021 | A multi-centric study on validation of the Fear Scale for COVID-19 in five Arabic speaking countries |
| Elagib et al [42] | 2022 | Knowledge, attitude, and practice regarding infection control measures among dental students during COVID-19 pandemic |
| Elhadi et al [43] | 2020 | The formidable task of fighting COVID-19 in Sudan |
| El-Sadig et al [44] | 2021 | Obstacles faced by healthcare providers during COVID-19 pandemic in Sudan |
| El-Sadig et al [45] | 2021 | Impact of COVID-19 on doctors and healthcare providers during the pandemic in Sudan |
| Fahal et al [46] | 2022 | The Mycetoma Research Centre experience during the COVID-19 pandemic: obstacles and beyond |
| Gayawan et al [47] | 2020 | The spatio-temporal pandemic dynamics of COVID-19 outbreak in Africa |
| Gilbert et al [48] | 2020 | Preparedness and vulnerability of African countries against importations of COVID-19: a modelling study |
| Gismalla et al<br>[49] | 2021 | Medical students' perception towards E-learning during COVID 19 pandemic in a high burden developing country |
| Hassan et al<br>[50] | 2020 | Convalescent plasma as a treatment modality for coronavirus disease 2019 in Sudan |
| Hezima et al<br>[51] | 2020 | Knowledge, attitudes, and practices of Sudanese residents towards COVID-19 |
| Lami et al [52] | 2022 | Risk Factors of COVID-19 Critical Outcomes in the Eastern Mediterranean Region: Multicountry Retrospective Study |
| Mahgoub et al<br>+ | 2021 | Psychological effects of the COVID-19 pandemic: Perceived stress, anxiety, work-family imbalance, and coping strategies among healthcare professionals in Khartoum state hospitals, Sudan |
| Mahmoud et al<br>[53] | 2021 | Awareness and perception of COVID-19 among final-year dental students, Sudan |
| Badawi et al<br>[54] | 2020 | Have We Passed the Peak? Prevalence and Clinical Presentations of Covid-19 among Sudanese Health Workers |
| Mehanna et al<br>[55] | 2021 | Public willingness to adhere to COVID-19 precautionary measures in Sudan: an application of the Health Belief Model |
| Mohamed et al<br>[56] | 2021 | Knowledge, attitude and practice of the Sudanese people towards COVID-19: an online survey |
| Moser et al [20] | 2022 | SARS-CoV-2 Antibody Prevalence and Population-Based Death Rates, Greater Omdurman, Sudan |
| Mukhtar &<br>Khogali [57] | 2021 | The accelerating COVID-19 pandemic in Sudan |
| Musa et al [58] | 2020 | COVID-19 in Sudan: Response towards control and prevention |
| Pigoga et al [59] | 2021 | Derivation of a Contextually-Appropriate COVID-19 Mortality Scale for Low-Resource Settings |
| Raja et al [60] | 2022 | COVID-19 vaccine acceptance, hesitancy, and associated factors among medical students in Sudan |
| Sayedahmed<br>et al [61] | 2020 | Knowledge, attitude and practice regarding COVID-19 among Sudanese population during the early days of the pandemic: Online cross-sectional survey |
| Siddig et al [62] | 2022 | COVID-19 and cognitive impairment: A cross-sectional clinic-based study |
| Tahir et al [63] | 2021 | Internet addiction and sleep quality among medical students during the COVID-19 pandemic: A multinational cross-sectional survey |
| Yassin et al [64] | 2022 | COVID-19 Vaccination Acceptance among Healthcare Staff in Sudan |
| Yousif et al [65] | 2021 | Use of Remdesivir in the treatment of Coronavirus Disease 2019 (COVID-19) infection among Sudanese patients: a case series |

### 3.4 Prevalence of pneumonia

Seven included studies determined prevalence of pneumonia. Three studies determined prevalence among suspected outpatients, one study determined prevalence among tuberculosis patients while the other three studies determine pneumonia prevalence among children. Furthermore, three studies determined prevalence of several microorganisms causing pneumonia; two studies were concerned with prevalence of *S. pneumonia*, one was concerned with prevalence of *M. Catarrhalis* and one was concerned with prevalence of *SARS-Cov-2*. Characteristics of studies are depicted in (Table 1).

The pooled prevalence of all seven studies using fixed effect model was 33.33% [CI: 33.32, 33.34] and 30.47% [CI: 10.41, 50.53] using the random effect model. Moreover, after conducting sensitivity analysis; fixed effect pooled prevalence was 30.02% [CI: 30.01, 30.03] and 26.45% [CI: 3.16, 49.74] using the random effect model. Heterogeneity was high among all Meta analysis conducted (I^2^=100%) (Fig 2).

**(Fig 2):**
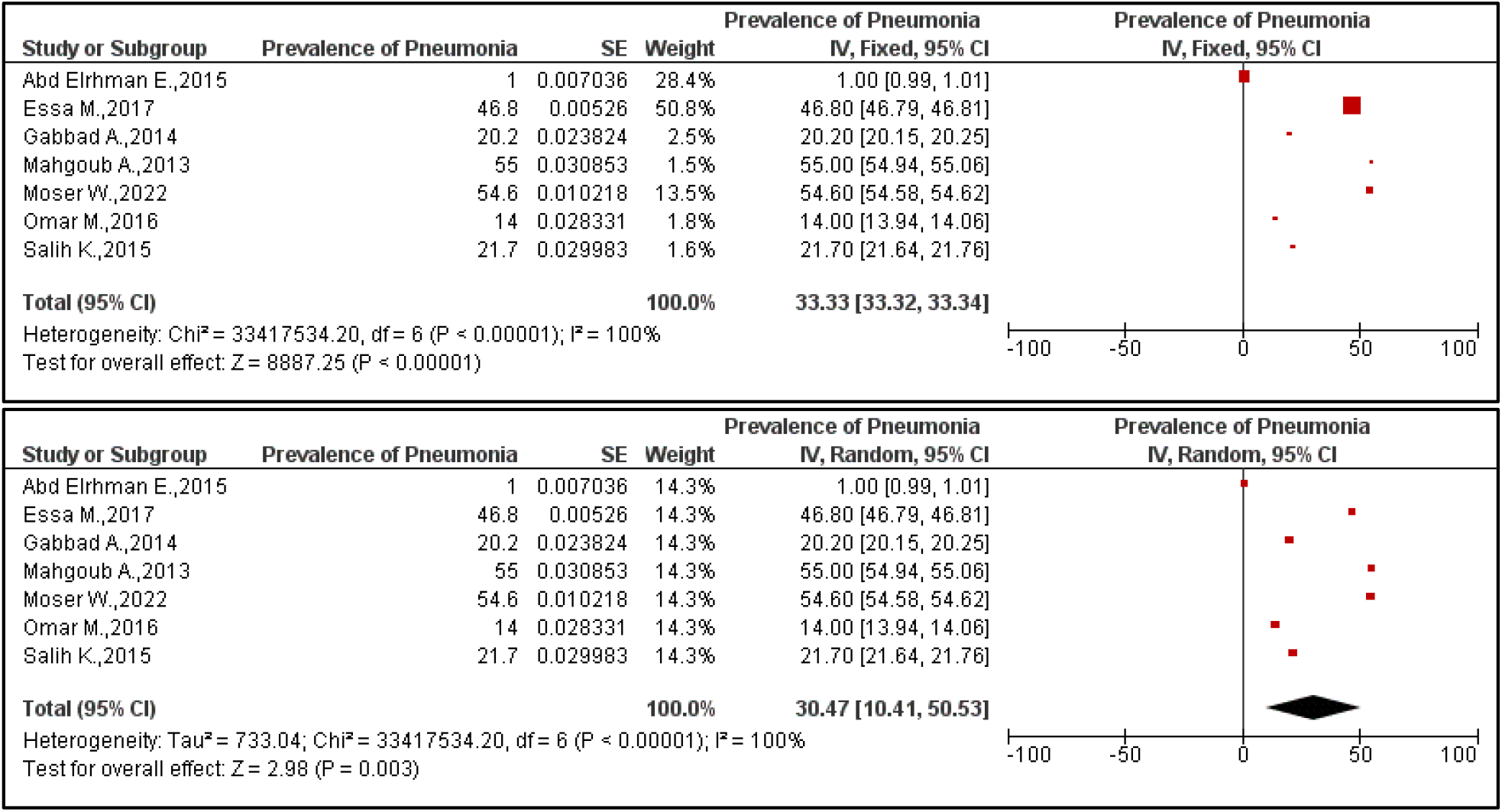
Pooled prevalence of pneumonia among included studies. **Above:** Fixed effect model. **Below:** Random effect model.

Children pneumonia prevalence were investigated among three included studies ^[14,17,25]^, pooled prevalence using fixed effect model was 29.94% [CI: 29.91, 29.97] and 32.30% [CI: 10.96, 53.64] using the random effect model (Fig 3).

**(Fig 3):**
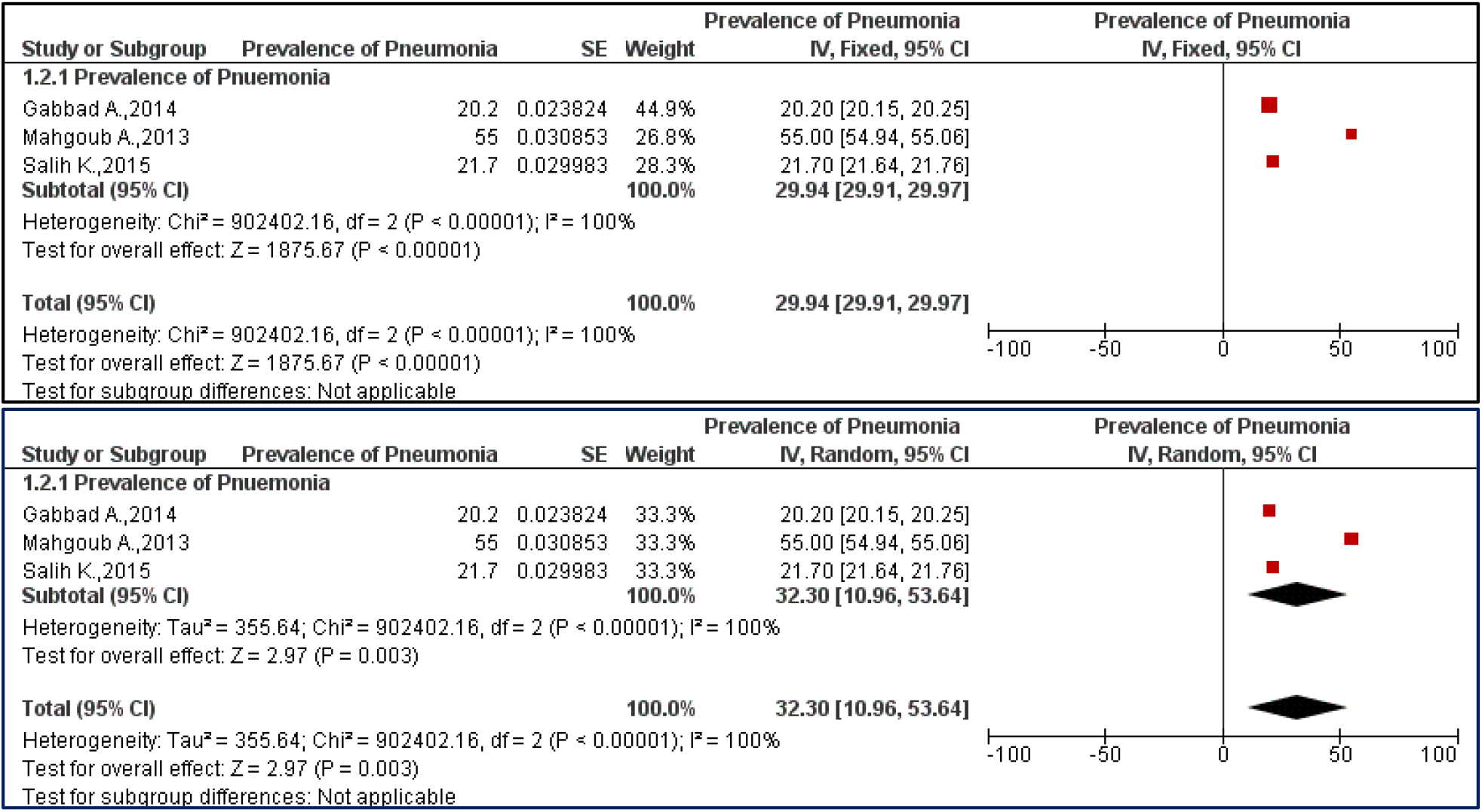
Pooled prevalence of children pneumonia among included studies. **Above:** Fixed effect model. **Below:** Random effect model.

### 3.5 Risk factors

Five research articles investigated risk factors related to pneumonia among Sudanese population.

The study of Gritly and colleagues ^[15]^ was conducted to determine the risk factors contributing to pneumonia at Omdurman city among 40 children under five years of age using a questionnaire, (57.50%) were males and less than one year, (67.50%) of patient’s fathers were labor (worker), (55.0%) with educations as basic, and (65.0%) of patient fathers make as less than 150 SDG (per day), moreover, most of the children (72.50%) of patients were breastfed. Majority of study participants (65.0%) used tap water, Most of the study participants (mothers) (47.50%) used *zeer* pot fridge for drinking water and storage No determination of any significant associations was described in the study.

The study of Salih and colleagues ^[23]^ investigated 224 children under 5 years of age who presented to the emergency units at Gaafar Ibn Oaf Children’s Hospital and Omdurman Children’s Hospital and were assessed and managed for severe presentation of pneumonia, the participants were (52.7%) female, (41.0%) less than one year of age, (29.0%) had 13-18 months breastfeeding, (65%) have unemployed parents, 67.9% were using Gas for cooking and 67.4% having father as a source of exposure to tobacco smoke. No determination of any significant associations was described.

Another study ^[14]^ was determined to measure the proportion of pneumonia among 282 children attending Omdurman Paediatric Hospital to highlight the disease related factors, using questionnaire. Their results indicated that males *OR* (2.41), mother illiteracy *OR* (7.54), low income *OR* (1.55), urban residence *OR* (2.09) are socio-cultural risks that are significantly related to children pneumonia.

Moreover, the study of Mahgoub and colleagues ^[17]^ was aimed to determine the prevalence of pneumonia among under ten years children in Khartoum State as well as to investigate associated factors using questionnaires. 400 families were included in their study. Income *OR* (2.73), mother education less than university *OR* (1.4), home size (1-2 rooms) *OR* (2.36) were concluded as socio-cultural risks that are significantly related to children pneumonia.

Furthermore, the study of Salih and colleagues ^[25]^ was aimed to determine risk factors of death among 195 children aged 2-59 months hospitalized for severe pneumonia in Khartoum State, authors investigated the significant change of outcome (recovery *vs* death) according to gender. Their result indicated that male are more likely to die *OR* (1.1).

### 3.6 Knowledge, practice and awareness assessment

Two research articles investigated Knowledge, practice or awareness related to pneumonia among Sudanese population.

A study ^[16]^ was aimed to assess nurses’ knowledge and practice regarding nursing care of patients with ventilator associated pneumonia using interview questionnaire at Ahmed Gasim Hospital in Khartoum State. Sixty nurses were included, only (34.5%) of participants responded correctly regarding signs of ventilator-associated pneumonia and prevention methods of ventilator associated pneumonia, (85%) correctly practiced gastric reflux prevention, (55%) practiced equipment maintenance, (73.69%) practiced suctioning from the ETT/tracheotomy, (68.65%) practiced care of patient with ventilator associated pneumonia. Moreover, another study ^[18]^ aimed to assess mothers’ knowledge and recognition of pneumonia among children less than 5 years of age in *bashir Banaga* village, Hassahisa locality, Gezira State as well as mothers’ attitude toward seeking medical help if they had a child with symptoms of pneumonia. 250 mothers were interviewed using questionnaire. Participants were (41.8%) 31-35 years old, 73.2% were unemployed. weather change (17.6%) and cold temperatures (12.4%) were indicated as causes. Only 26 % of mothers reported that a virus or germ causes pneumonia. Moreover, only (23.6%) of mothers said they would recognize pneumonia if their child had rapid breathing or (34.0%) if the child had cough.

The study of Mohamoud ^[19]^ was aimed to investigate Knowledge, Attitude and Practice of 182 parents of children suffered from pneumonia less than five years of age in Daraga district, Wad Madani town, Gezira State using questionnaires. 81.4% of parents were 30-49 years of age, 70.3% were still married, 38.5% have secondary education as the high degree, and 44.0% were housewives. Moreover, (70.3%) of parents said Virus causes pneumonia in children, (23.1%) said bacteria, (2.2%) listed congenital, (2.7%) listed parasites, and (1.6%) said other. Furthermore, (31.3%) of the parents listed fever and cough as symptoms of pneumonia, (13.7%) listed chest pain, (3.3%) listed vomiting, (51.1%) listed fever cough and chest pain and (0.5%) listed other. As routes of transmission (28.0%) of the parents said coughing, while (31.9%) said sneezing, (1.6%) said handshake, and (37.4%) said all of them. As prevention strategy, (59.9%) of the parents listed vaccination, good health and complete cycle of breastfeeding to the child as best ways to prevent childhood pneumonia, Most of parents (73.6%) indicated that they had no animals’ shelter in their houses.

### 3.7 Other outcomes

The study of Salah and colleagues ^[22]^ was aimed to determine the prevalence of hypoxemia among less than five years children with pneumonia with presented with an acute history of cough and rapid respiration or difficulty in breathing in Omdurman city, Khartoum State. Out of 150 children participated, (57.3%) were males, (32%) were 2 to ≤ 12 months. 42.7% of patients had hypoxemia (with pulse oximeter oxygen saturation < 90%), 36 (56.25%) were in the age group < 2 months. Of the hypoxic patients, 30 (46.88%) have severe pneumonia and 7 (10.94) have very severe pneumonia. Moreover, Hypoxemia also increased in male sex but not of statistical significance (*P* = 0.72) and Hypoxemia significantly increased in patients diagnosed as very severe disease (*P* < 0.001).

The study of Salih and colleagues ^[24]^ was designed to investigate the adherence and response of the *WHO* guidelines for treatment of severe pneumonia in Khartoum State. 208 children aged 2 to 59 months with definite labeling diagnosis of “severe pneumonia” were included. The mean (SD) of the age was reported as 28.12 (13.9) months and (52.4%) were females. Only 39 (18.8%) of these children received the prescription that was adherent to the *WHO* guidelines (penicillin) of severe pneumonia. The antimicrobial prescriptions, which were not in accordance with the *WHO* guidelines, were: amoxicillin/ clavulanic acid (46, 22.1%), ceftriaxone (42, 20.2%), cefuroxime (41, 19.7%), a combination of penicillin/gentamicin (29; 13.9%) and others 11 (11, 5.3%). None of the investigated factors (age, gender, symptoms and signs) were found as predictors for adherence of the *WHO* guidelines for treatment of severe pneumonia. There was no significant difference in the response between adherence and non-adherence prescriptions. Lastly, none of the investigated predictors was found to be associated with treatment outcome

## 4 Discussion

To our knowledge, this scoping review is the first attempt to find out the magnitude of information on pooled prevalence of pneumonia as well as its associated risk factors in Sudan. A widespread search from several published databases and stringent methodology to screen and include every potential study was approached in the present study.

Pooled prevalence of pneumonia among Sudanese children was around 30% considering the differences between fixed and random effect models, lower estimates have been reported in Uganda (25%), Kenya (21%). However, an almost similar result has been reported in Ethiopia (33%) ^[66,67]^.

Several studies among included assessed socio-cultural risk factors related to acquisition of pneumonia; with male gender, illiteracy, low income and urban residence as the main socio-cultural related factors. This is similar to what had been reported that gender distribution indicated that pneumonia was more prevalent in male than in female in previous several reports ^[68–70]^.

Moreover, a study conducted in Uganda ^[67]^ indicated that children of rural residence had 5.7 higher odd of having pneumonia compared to the urban residence. Rural residence reported in Uganda finding (OR=5.7, 95%CI=2.97-11.05, p <0.001), is comparable with a study conducted in Ethiopia as well ^[71]^. which reported 4.5 higher odds of developing pneumonia among children of rural residence. Nevertheless, due to exposure to factors as air pollution and overcrowding in urban areas other studies indicated that the odds are higher in urban compared to rural. Further research is needed to investigate specific ecological risk factors to better study the urban/rural pneumonia infection patterns^[72]^.

Furthermore, it is concluded that incidence of pneumonia is higher in people at the extremes of age as well as people living in socially deprived areas as indicated by several studies, this finding is in agreement with several research articles in literature ^[73,74]^.

Regarding socio-economic status and supporting findings from included studies; a study conducted by Park in 2007 reported that children from low socio-economic status tend to have more risk of respiratory infections and that children of mothers with a personal source of income are at a lower risk of developing pneumonia as indicated by a study conducted in Gambia by O’Dempsey and colleagues ^[5,75–77]^.

Furthermore, (43.5%) of nurses in the study of Idress ^[16]^ was correctly responded regarding signs of ventilator-associated pneumonia and prevention methods of ventilator associated pneumonia, this is similar to what had been reported in USA by study of Mietto and colleagues in 2013 where average knowledge score regarding signs and symptoms of ventilator-associated pneumonia was (43.28)%. Moreover, poor knowledge had been reported as more than 65% in Sudan in a study by Elbokhary & Osama in 2015 ^[78,79]^.

This scoping review identified several key gaps within the existing body of literature. Studies concerning pneumonia in Sudan are generally scarce that Meta analysis cannot be synthesized. Moreover, no studies – to our knowledge were conducted to investigate specific ecological risk factors which may facilitate pneumonia rather than “rural residence”. Moreover, the risk of pneumonia is partially driven by host genetics. *CYP1A1* for example is a widely studied pulmonary *CYP* family gene primarily expressed in peripheral airway epithelium. No studies – to our knowledge were conducted to investigate related genetic variations of infected participants and its possible relations to pneumonia or its severity.

### Strengths and Limitations

The strengths of this review are that we systematically identified and included related studies from 2010 to 2020, Moreover; we have conducted meta-analysis to derive pooled prevalence estimates of studies related. Furthermore, we carried out a quality assessment of the included studies based on criteria specifically developed to determine the quality of included studies.

Nevertheless, several limitations are to be considered when interpreting study results; grey literature evidence was not assessed. Moreover, African journals that are not indexed in the screened databases was not considered for inclusion as well, although all included studies are of good quality, several good studies might have been missed. Lastly, the heterogeneity was high among the Meta analysis conducted.

## Conclusion

The current study findings indicate that the pooled prevalence of pneumonia is around 30% for general population as well as children. Further research with larger sample sizes targeting prevalence and risk factors of pneumonia among Sudanese population is needed to be conducted.

## Data Availability

All data produced in the present work are contained in the manuscript

### Appendices

**Table (S1):**
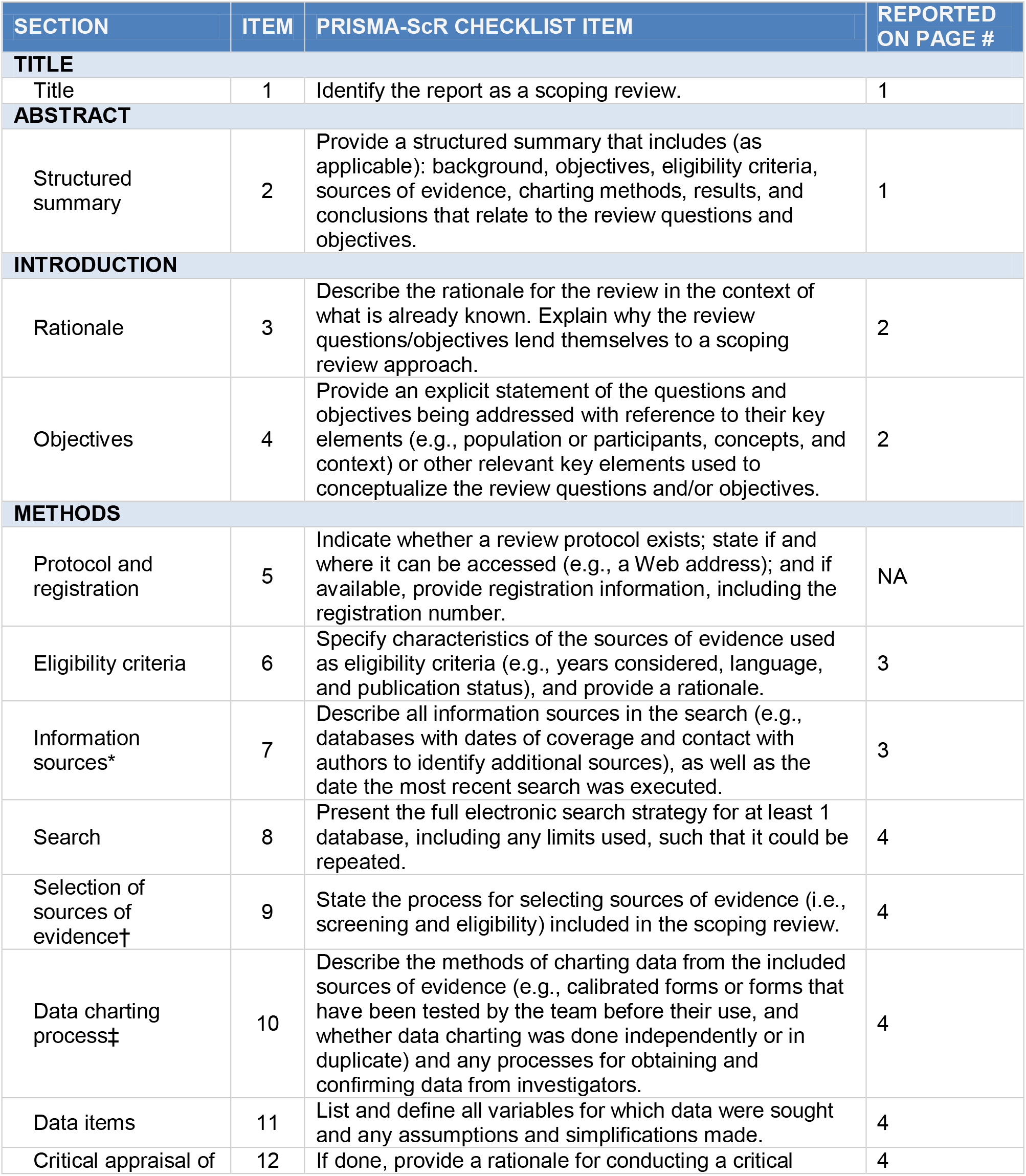
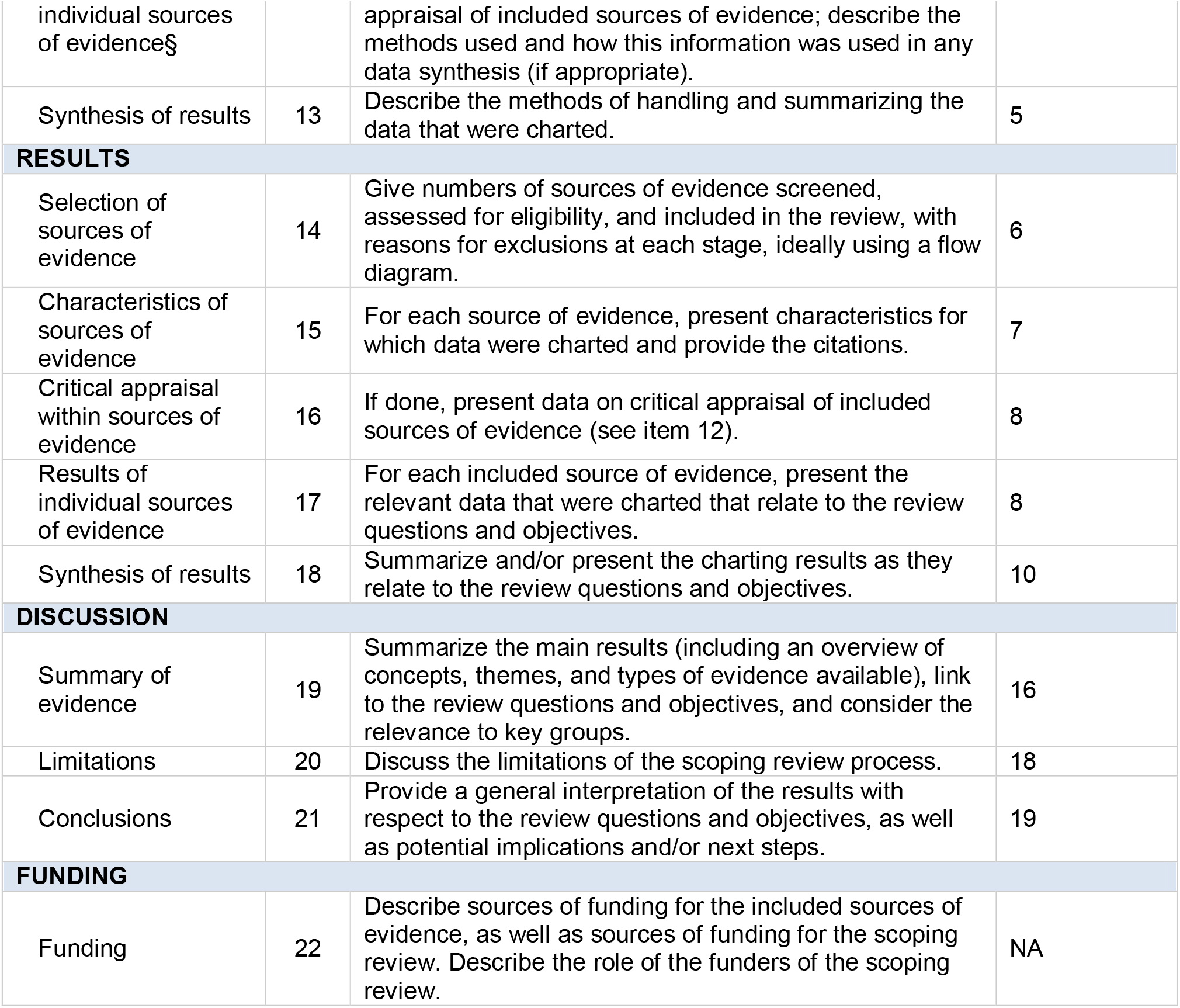
Preferred Reporting Items for Systematic reviews and Meta-Analyses extension for Scoping Reviews (PRISMA-ScR) Checklist

| SECTION | ITEM | PRISMA-ScR CHECKLIST ITEM | REPORTED ON PAGE # |
| --- | --- | --- | --- |
| **TITLE** |  |  |  |
| Title | 1 | Identify the report as a scoping review. | 1 |
| **ABSTRACT** |  |  |  |
| Structured summary | 2 | Provide a structured summary that includes (as applicable): background, objectives, eligibility criteria, sources of evidence, charting methods, results, and conclusions that relate to the review questions and objectives. | 1 |
| **INTRODUCTION** |  |  |  |
| Rationale | 3 | Describe the rationale for the review in the context of what is already known. Explain why the review questions/objectives lend themselves to a scoping review approach. | 2 |
| Objectives | 4 | Provide an explicit statement of the questions and objectives being addressed with reference to their key elements (e.g., population or participants, concepts, and context) or other relevant key elements used to conceptualize the review questions and/or objectives. | 2 |
| **METHODS** |  |  |  |
| Protocol and registration | 5 | Indicate whether a review protocol exists; state if and where it can be accessed (e.g., a Web address); and if available, provide registration information, including the registration number. | NA |
| Eligibility criteria | 6 | Specify characteristics of the sources of evidence used as eligibility criteria (e.g., years considered, language, and publication status), and provide a rationale. | 3 |
| Information sources* | 7 | Describe all information sources in the search (e.g., databases with dates of coverage and contact with authors to identify additional sources), as well as the date the most recent search was executed. | 3 |
| Search | 8 | Present the full electronic search strategy for at least 1 database, including any limits used, such that it could be repeated. | 4 |
| Selection of sources of evidence† | 9 | State the process for selecting sources of evidence (i.e., screening and eligibility) included in the scoping review. | 4 |
| Data charting process‡ | 10 | Describe the methods of charting data from the included sources of evidence (e.g., calibrated forms or forms that have been tested by the team before their use, and whether data charting was done independently or in duplicate) and any processes for obtaining and confirming data from investigators. | 4 |
| Data items | 11 | List and define all variables for which data were sought and any assumptions and simplifications made. | 4 |
| Critical appraisal of | 12 | If done, provide a rationale for conducting a critical | 4 |
| individual sources of evidence§ |  | appraisal of included sources of evidence; describe the methods used and how this information was used in any data synthesis (if appropriate). |  |
| Synthesis of results | 13 | Describe the methods of handling and summarizing the data that were charted. | 5 |
| **RESULTS** |  |  |  |
| Selection of sources of evidence | 14 | Give numbers of sources of evidence screened, assessed for eligibility, and included in the review, with reasons for exclusions at each stage, ideally using a flow diagram. | 6 |
| Characteristics of sources of evidence | 15 | For each source of evidence, present characteristics for which data were charted and provide the citations. | 7 |
| Critical appraisal within sources of evidence | 16 | If done, present data on critical appraisal of included sources of evidence (see item 12). | 8 |
| Results of individual sources of evidence | 17 | For each included source of evidence, present the relevant data that were charted that relate to the review questions and objectives. | 8 |
| Synthesis of results | 18 | Summarize and/or present the charting results as they relate to the review questions and objectives. | 10 |
| **DISCUSSION** |  |  |  |
| Summary of evidence | 19 | Summarize the main results (including an overview of concepts, themes, and types of evidence available), link to the review questions and objectives, and consider the relevance to key groups. | 16 |
| Limitations | 20 | Discuss the limitations of the scoping review process. | 18 |
| Conclusions | 21 | Provide a general interpretation of the results with respect to the review questions and objectives, as well as potential implications and/or next steps. | 19 |
| **FUNDING** |  |  |  |
| Funding | 22 | Describe sources of funding for the included sources of evidence, as well as sources of funding for the scoping review. Describe the role of the funders of the scoping review. | NA |

**Table (S2):** Quality assessment of included studies

| **Study ID** | **Is the study objective clearly defined** | **Is the study sample completely determined** | **Is the study population clearly defined and specified** | **Is the methodology rigorous** | **Is the data analysis rigorous** | **Final score** |
| --- | --- | --- | --- | --- | --- | --- |
| Abd Elrhman E.,2015 | 1 | 1 | 1 | 1 | 0 | 4 |
| Essa M.,2017 | 1 | 1 | 1 | 1 | 0 | 4 |
| Gabbad A.,2014 | 1 | 1 | 1 | 1 | 1 | 5 |
| Gritly S.,2018 | 1 | 1 | 1 | 1 | 1 | 5 |
| Idress W.,2017 | 1 | 1 | 1 | 1 | 1 | 5 |
| Mahgoub A.,2013 | 1 | 1 | 1 | 1 | 1 | 5 |
| Mohammed S.,2013 | 1 | 1 | 1 | 1 | 1 | 5 |
| Mohamoud A.,2014 | 1 | 1 | 1 | 1 | 1 | 5 |
| Moser W.,2022 | 1 | 1 | 1 | 1 | 1 | 5 |
| Omar M.,2016 | 1 | 1 | 1 | 1 | 1 | 5 |
| Salah E.,2015 | 1 | 1 | 1 | 1 | 1 | 5 |
| Salih K.,2011 | 1 | 1 | 1 | 1 | 1 | 5 |
| Salih K.,2014 | 1 | 1 | 1 | 1 | 1 | 5 |
| Salih K.,2015 | 1 | 1 | 1 | 1 | 1 | 5 |
| Salih K.,2015 | 1 | 1 | 1 | 1 | 1 | 5 |

